# The Maudsley 3-item visual analogue scale (M3VAS): Longitudinal validation of a new measure to capture depression

**DOI:** 10.1101/2023.06.26.23291655

**Authors:** Maria Elena Middag, Daniel Silman, Anthony Cleare, Allan Young, Ben Carter, Rebecca Strawbridge

## Abstract

The existing ‘gold standard’ assessments of depression suffer various issues, which include the evaluation of constructs extraneous to the core symptoms of the illness, and low sensitivity to change over time. Members of our team developed a new, simple visual analogue scale that aims to address these issues (M3VAS). Initial validation of the scale demonstrated good internal consistency and convergent validity. Still, we have not yet investigated how well it assesses changes in the severity of depressive symptoms over time. This project will analyse data from a longitudinal study (Hampsey et al., 2022), aiming to ascertain whether the M3VAS is sensitive to change i.e., how well it detects either a worsening or improvement of symptoms over a number of weeks (repeated measures). Validating this scale’s longitudinal validity could provide a better way for future longitudinal (observational and interventional) studies to identify changes in the severity of depressive illness.

## Introduction

While there are several assessments used to assess the severity of depressive symptoms, the existing gold standard measures, such as the Hamilton Rating Scale for depression (HAMD) (Hamilton, 1967), the Montgomery Åsberg Depression Rating Scale (MADRS) (Montgomery & Åsberg, 1979), the Inventory of Depressive Symptomatology (IDS) (Rush et al., 1986) and Patient Health Questionnaire (PHQ-9) (Kroenke et al., 2001), possess some limitations, including evaluating constructs extraneous to the core symptoms of depression and low sensitivity to change over time (Carrozzino, Patierno, Fava & Guidi, 2020). Thus, there is a need for a new assessment that can sensitively measure changes in the severity of core depressive symptoms over time.

To address this need, the M3VAS was developed as a simple visual analogue scale that aims to measure depression severity without evaluating constructs extraneous to the core symptoms of depression. Initial validation of the M3VAS demonstrated good internal consistency including a single-factor model and convergent validity with the 16-item Quick Inventory of Depressive Symptomatology (QIDS) (Moulton et al., 2021). Although the M3VAS is a validated tool for measuring depression, the longitudinal validity of the M3VAS has not yet been established.

### Hypotheses

H_1_: The M3VAS can reliably measure changes in depressive symptoms over **2 weeks** compared to Day 0 (baseline).

H_2_: The M3VAS can reliably measure changes in depressive symptoms over **4 weeks** from week 2 and Day 0.

H_3_: There will be a positive correlation between the M3VAS and PHQ-9 continuous severity scores.

### Study Objectives

This study’s primary objective is to investigate the sensitivity of the M3VAS in detecting changes in depressive symptoms over time. It is hoped that this will provide a better way for future longitudinal studies to identify changes in the severity of depression (particularly in ascertaining the efficacy of interventions and detecting relapse).

## Methods

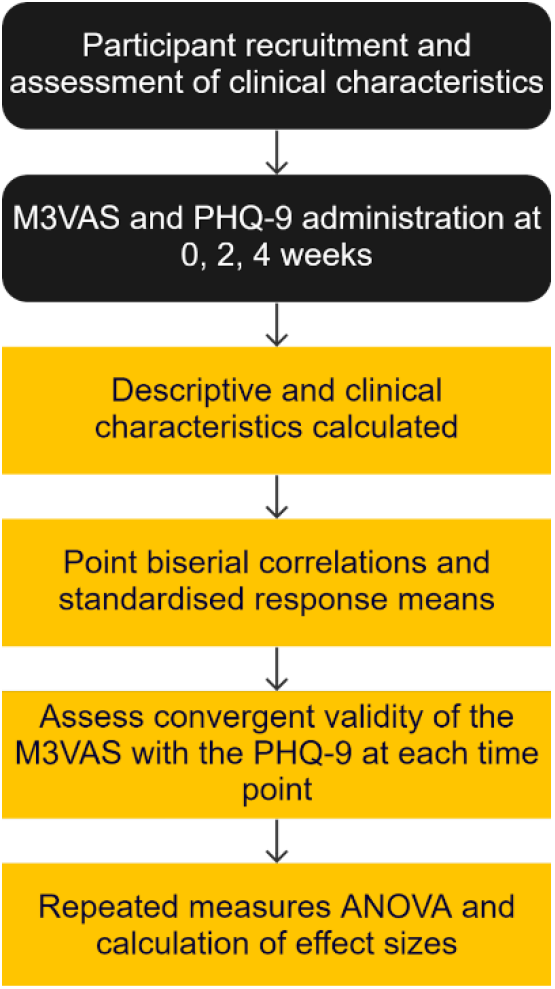

### Design

The study will analyse the data collected from the “Rhapsody” study (Hampsey et al., 2022) concerning the longitudinal validity of the M3VAS in detecting changes in core depressive symptoms and suicidality over time. This study was given ethical approval (REC reference: 21/ PR/0070) in April 2021. All participants completed assessments, including the Patient Health Questionnaire-9 item (PHQ-9) and the M3VAS, at three time points: baseline (Day 0), 2 weeks, and 4 weeks.

At the time of writing this protocol, data are available but have not yet been accessed.

### Participants

#### Affective disorders group

Participants with affective disorders are aged 18–85 years, diagnosed with either unipolar major depression (n=25) or bipolar disorder (n=25), and in a current depressive episode according to the Diagnostic and Statistical Manual of Mental Disorders, fourth edition (DSM-IV) criteria.

#### Unaffected matched controls group

Healthy controls were matched for gender, age and education levels (n=26). Participants in this group did not meet criteria for a depression diagnosis, were judged to be in good health although were permitted to experience mild disorders that do not impair daily functioning.

#### Eligibility criteria

The study will include the affective disorders (depression) group collected by Hampsey et al. (2022) and the matching healthy control group.

### Measures

All participants were asked to complete the PHQ-9 and the M3VAS at each timepoint (week 0, 2, 4). Participants in the depression group were assessed by the Mini-International Neuropsychiatric Interview (M.I.N.I.) V.5.0 (Sheehan et al., 1998), with at least moderate severity as assessed on the Clinical Global Impressions Scale (CGI) (Guy, 1976). The M.I.N.I. suicide questionnaire scale was only completed by participants scoring over 9 on the PHQ-9.

## Data Analysis

The present study will examine whether the M3VAS is sensitive to change over time. It will do this by comparison of what depression change looks like over time as measured by the PHQ-9. To examine the convergent validity of the M3VAS with the PHQ-9, Pearson’s correlation coefficients will be calculated to assess the convergent validity of the M3VAS with the PHQ-9 at each time point for the depressed participants and control patients combined.

Descriptive statistics such as the standard deviation of the M3VAS scores at each time point separately will be calculated for both groups and presented visually with Boxplots. Profile plots of the individual mean changes over time will be plotted, with the associated SD. As well as this, standardised response means will be calculated to account for different levels of variance in the data at baseline and follow-up. This will be done at baseline, 2, and 4 weeks for both the PHQ-9 and the M3VAS. The standardised response mean (mean change/SD_mean change_) will be estimated to show a simple reported co-efficient of change for each summary.

We will present the correlation of the M3VAS change compared to the PHQ-9 depression change severity over time to show the test-retest reliability across weeks. This will be measured both across and within participants.

To further evaluate the M3VAS’s longitudinal validity, a Bland-Altman analysis will be conducted to assess agreement between the M3VAS and PHQ-9 scores at each time point. This will be shown visually on a Bland-Altman plot to provide insight into the variability between the measures. Mean difference and limits of agreement will be calculated, representing the expected range of differences between the two measures.

A repeated measures ANOVA will be conducted to examine the main effect of time on M3VAS scores for each group separately (i.e., depressed and healthy), as well as the interaction effect of time and group. Time (0, 2, and 4 weeks) will be the within-subjects factor, and group (depressed and healthy) will be the between-subjects factor. Effect sizes will be calculated using partial eta squared. This will be to see if there are significant changes in M3VAS scores over time and whether these changes differ between the depressed and healthy groups.

The rates of missing data will be described for each depression severity measure at each time point. We will describe both the proportion of patients who didn’t complete the measures at all or partially didn’t complete the items in the measures.

### Statistical software

All statistical analyses will be conducted using the SPSS statistical software package.

## Data Availability

No data are available.

## Data Availability

All data produced in this study will be available upon reasonable request to the authors.

## Funding & Acknowledgements

This research is supported by the National Institute for Health and Care Research (NIHR) Maudsley Biomedical Research Centre in South London, the Maudsley NHS Foundation Trust and King’s College London. The views expressed are those of the authors and not necessarily those of the NIHR or the Department of Health and Social Care. The authors note that the content of this manuscript has not been published or submitted for publication elsewhere. The authors express their sincere gratitude to the company Novoic who sponsored the study, and the particular involvement of Caroline Skirrow, Marton Meszaros, and Emil Fristed from Novoic. We also express our sincere gratitude to Rosie Taylor and Elliot Hampsey who coordinated all recruitment and data collection, and thanks also to our study participants. Project RHAPSODY was funded by the National Institute of Health and Care Research (NIHR) and NHSX; AI_AWARD01984.

